# Development and implementation of a clinical decision support system tool for the evaluation of suspected monkeypox infection

**DOI:** 10.1101/2022.08.15.22278791

**Authors:** John S. Albin, Jacob E Lazarus, Kristen M. Hysell, David M. Rubins, Lindsay Germaine, Caitlin M. Dugdale, Howard M. Heller, Elizabeth L. Hohmann, Joshua J. Baugh, Erica S. Shenoy

## Abstract

Monkeypox virus was historically rare outside of West and Central Africa until the current 2022 global outbreak, which has required clinicians to be alert to identify individuals with possible monkeypox, institute isolation, and take appropriate next steps in evaluation and management. Clinical decision support systems (CDSS), which have been shown to improve adherence to clinical guidelines, can support frontline clinicians in applying the most current evaluation and management guidance in the setting of an emerging infectious diseases outbreak when those guidelines are evolving over time. Here we describe the rapid development and implementation of a CDSS tool embedded in the electronic health record to guide frontline clinicians in the diagnostic evaluation of monkeypox infection and triage patients with potential monkeypox infection to individualized infectious diseases physician review. We also present data on the initial performance of this tool in a large integrated healthcare system.

## INTRODUCTION

In early May 2022, a cluster of monkeypox cases was initially identified in Great Britain, with the first identified case in the United States later that month.^1^ The extent of the global monkeypox outbreak has continued to spread since, affecting tens of thousands of individuals more than 80 non-endemic countries by mid-August 2022.^2^ Prompt recognition of monkeypox infection is important not only to expedite potential initiation of therapy, but also to institute transmission-based precautions to minimize risks of monkeypox transmission within healthcare facilities. As knowledge about the epidemiological risk factors and clinical presentation of people with monkeypox infection has grown during this outbreak, the definition of a person under investigation (PUI), a key element in driving diagnostic evaluation, has evolved, as have testing and infection control protocols. This rapidly evolving knowledge necessitates prompt dissemination of information to frontline clinicians as well as iterative adaptation of clinical resources to remain up to date with dynamic local and national guidelines.

Clinical decision support systems (CDSS), especially those computerized products that provide automated decision support as part of clinician workflow, have been shown to improve adherence to guidelines.^3,4^ In our large integrated health system, two years after the initial development of a COVID-19 CDSS, isolation, testing, and de-isolation for patients with suspected and confirmed COVID-19, is largely automated, with only a small subset of the most complex cases triaged to clinical or infection control review.^5^ Building on this experience, we sought to create a CDSS to assist frontline clinicians in the evaluation of patients presenting with symptoms concerning for monkeypox. Here, we describe the development of the Monkeypox (MPX) Clinical Decision Support Tool, a dynamic CDSS embedded in the electronic health record (EHR) and capable of adapting to the outbreak as it develops. We detail major innovations and provide initial outcomes of monkeypox PUIs evaluated during early deployment

## METHODS

Massachusetts General Hospital (MGH) is a large academic medical center in Boston, MA that was the site of the first case of confirmed monkeypox infection in the US on May 18, 2022.^1^ Immediately following that first diagnosis, frontline clinicians were instructed to contact a team of infectious disease physicians (i.e., the MGH Biothreats Team) who have expertise in the evaluation of high-consequences infectious diseases when patients presented for care with a new rash and epidemiologic risk factors for monkeypox infection. Biothreats team physicians then individually reviewed patient details with the frontline clinicians and facilitated additional diagnostic workup for monkeypox at the Massachusetts Department of Public Health, when indicated.

Informed by these early evaluations of potential monkeypox PUIs, and based on prior experience in developing and implementing CDSS for COVID-19, Biothreats team physicians collaborated with MGH infection control leadership and members of the Mass General Brigham (MGB) Digital Health Team to develop and implement the MPX Clinical Decision Support Tool into the healthcare system’s electronic health record (Epic Systems Inc., Verona, WI). MGB is a large integrated healthcare system in Massachusetts and New Hampshire which includes MGH, as well as eight acute care hospitals, two specialty hospitals, and a broad network of post-acute care, community health, and primary care centers. Across the system, the MPX tool was launched as an on-demand template that could be accessed by clinicians by typing “.MONKEYPOX” in any inpatient or outpatient note field.

The MPX CDSS tool consisted of a structured set of screening questions adapted from U.S. Centers for Disease Control and Prevention (CDC) criteria for establishing PUI status for monkeypox infection.^6^ Based on a clinician’s answers to questions presented, if a patient met both epidemiologic and clinical criteria for MPX PUI status, a secondary set of questions developed by Biothreats Team members were automatically added to the note to prompt the clinician to capture more data on individualized epidemiological risk for each patient. These questions included risk related both to the current monkeypox outbreak and to the classical modes of monkeypox transmission. A detailed description of the cascading questions and prompts are provided (**Supplement, Figures 1-11**).

**Figure 1.** Care settings in which the MPX CDSS tool was used. The distribution of care settings in which the tool was utilized during its first 6 weeks of implementation are shown. MPX: monkeypox; ID: infectious diseases.

Clinicians were additionally prompted in the MPX CDSS tool to upload images of the patient’s rash into the EHR to aid in virtual review by infectious disease and public health officials. The tool also included directions regarding immediate infection control protocols. Finally, to ensure the outcome of the PUI assessment was communicated across clinicians caring for the patient, the tool automatically added a label, called an infection status, to the patient record to indicate they were a PUI for monkeypox (“MPX-Risk”) upon completion of the note. The MPX-Risk infection status, similar to other infection statuses used to communicate the need for specific transmission-based precautions, can be seen by all users of the EHR and drives other automated systems (e.g., isolation orders) as well as feeding into data and analytics across the EHR.

On 6/8/2022, the MPX CDSS tool was launched across MGB with the ultimate goal of standardizing evaluation and management of monkeypox PUIs throughout the system. At each site, once the clinician completed the note, facility-specific instructions on the next point of contact (i.e, infection control, infectious diseases, etc), was provided based on facility-specific workflows. We evaluated outcomes between 6/8/2022-7/20/2022 including overall utilization, determination of PUI status, testing frequency, and test positivity among patients evaluated with the MPX CDSS tool. Records for patients seen in the MGH Sexual Health Clinic, subject to 42CFR protection, were excluded from this analysis. Over the course of the study period, several updates were made to capture the evolving PUI definition, collect information to facilitate decisions regarding home isolation, and modifications for end-user clarity.

This study was conducted under Mass General Brigham IRB protocol 2012P002359.

## RESULTS

### MPX Tool utilization

Between 6/8/2022-7/20/2022, the MPX CDSS tool was used for 55 distinct patients. Utilization within the MGB system occurred predominantly within the MGH with 39 encounters. Other MGB system evaluations included 5 at Brigham & Women’s Hospital (BWH), 5 at Newton-Wellesley Hospital (NWH), 1 at Cooley-Dickinson Hospital (CDH), 1 at Salem Hospital (SLM), and 4 across several urgent care settings. More than half of these completed encounters took place in an Emergency Medicine context (37), with most of the remainder occurring either in primary care settings (8) or the MGH infectious diseases clinic (6). The predominant use of the tool at MGH reflects overall volume of distribution of PUIs across the MGB healthcare system, which was focused at MGH (**Figure 1**).

### Descriptive characteristics of PUIs

The CDC PUI definition used to identify PUIs is provided (**Supplement Table 1**). Among 55 patients presenting with an unexplained rash for which the tool was used, 31 (56%) were identified by the tool to be possible PUIs. High rates of PUI identification were seen in the MGH Infectious Diseases Clinic (5 PUIs identified among 6 uses of the MPX tool, 83%). The PUI identification rate in an Emergency Medicine setting was 16/37 (43%), while 3/8 (38%) patients in primary care settings were determined to be possible PUIs for monkeypox. The average age among possible PUIs was 34.7 (SD 8.6) compared to 41.0 (SD 15.1) for patients who were not identified as possible PUIs. 26 possible PUIs identified as men, one as a transgender man, and 4 as women.

All patients identified as possible PUI, by definition, met the clinical PUI criterion of having an otherwise unexplained rash. Among the 31 patients who also met epidemiological criteria and thus were identified by the MPX CDSS tool as possible PUIs for monkeypox, 24 reported close contact with members of a social network experiencing monkeypox, 7 reported contact with a person with a similar rash or who was thought to have monkeypox, and 2 reported travel to an area of active monkeypox transmission. No patients reported contact with animals or animal products as described with monkeypox transmission in endemic settings.

### Testing outcomes

Among the 31 patients identified as possible PUIs by the MPX CDSS tool, testing was not pursued for 7 after discussion with the specialist. These patients were assessed as low risk on specialist review either at the hospital or in discussion with the state epidemiologist. Of the 24 patients for whom monkeypox testing was pursued, 5 (21%) were PCR positive, and 15 (63%) were PCR negative. 2 (8%) patients had inconclusive tests results, generally indicating a lack of amplification from the assay positive control; neither of these was retested. Results from 1 patient (4%) were not recorded in the medical record, and 1 patient (4%) was lost to follow-up prior to testing. We are aware of an additional 3 patients who tested positive for monkeypox at MGH during this period who were not triaged through the MPX tool; all presented through the infectious diseases clinic. The average age of patients who tested positive for MPX was 33.9 (SD 8.5) years; all were male. A summary of the MPX CDSS tool utilization and downstream outcomes by week over the first 6 weeks of implementation is presented (**Figure 2)**.

**Figure 2.** Trends in use of the MPX CDSS Tool. Bars depict the number of patients for whom the tool was used (total use), the number of possible PUIs identified by the tool (CDSS PUI)), the number of possible PUIs in whom testing was pursued after expert review (Testing Pursued), and the number of patients who tested positive (MPX positive) during each of the first six weeks of implementation. PUI: person under investigation; CDSS: clinical decision support system; MPX: monkeypox.

### Versioning

During the study period, updates occurred as expected to respond to changing epidemiology, infection control protocols, as well as end-user feedback to improve utilization. While the tool can be added to individual clinician’s notes or their personal library of note templates, it was constructed as a central system template in the EHR to maintain consistency across users. As updates were requested by the Biothreats Team to adapt to state or national guidance or changes in the epidemiology of the outbreak, these changes were automatically pushed to all users of the note simultaneously (**Figure 3**).

**Figure 3.** Versioning of the MPX CDSS Tool. The timeline showing major milestones prompting development of the MPX CDSS Tool (e.g., first diagnosed case at MGH on 5/18/2022), followed by request for development and implementation within 24 hours of request on 6/8/2022. The top half of the timeline describes multiple changes to the PUI definition and IC protocols, while the bottom half describes versioning related to changes in the CDS to allow for more efficient collection of pertinent patient information, as well as ensuring access by clinicians to the most up to date public health guidance and local points of contact. MPX: monkeypox; CDSS: clinical decision support system; CDC: Centers for Disease Control and Prevention; PUI: person under investigation; IC: infection control.

## DISCUSSION

We have described the development, implementation, and early usage patterns of an embedded clinical decision support system to facilitate the initial isolation and evaluation of patients in whom monkeypox infection is suspected. Once the decision was made to develop this CDSS, we were able to implement the MPX CDSS tool across the MGB system in a matter of days, and over the first 6 weeks, this tool was utilized 55 times for the evaluation of monkeypox infection in our healthcare system.

Importantly, this CDSS allows for rapid and frequent iterations with input of subject matter experts based on evolving epidemiological risk factors, testing strategies, and identified areas to improve the efficiency of diagnostic evaluation. Improvements such as automated patient chart labeling to identify patients who are PUIs and thus alert clinicians of the monkeypox work up in progress, and the ability to update front-line clinicians of infection control protocols in real-time, have provided the flexibility required in an emerging outbreak.

Several limitations are noted. The use of the MPX CDSS tool requires clinician buy-in as there is no forcing function within the EHR to compel use, and this can lead to missed opportunities for tool application. Despite this, with messaging to frontline clinicians and attention to integration into standard workflow, clinician acceptance of the tool can be achieved. In this study, we did not aim to evaluate the sensitivity or specificity of a the MPX CDSS as a clinical decision tool; MPX CDSS tool questions were derived from CDC definitions, but more data will be needed to validate the test characteristics of the instrument.

This approach has facilitated up-to-date decision support to frontline providers by centralizing MPX tool updating within a team of subject matter experts, including feedback from end users to improve functionality. Though described here only for monkeypox, developing similar tools for the evaluation of other high consequence infectious diseases such as Middle East Respiratory Syndrome (MERS), Ebola Virus Disease, and Avian Influenza may also help to facilitate efficient and adaptable evaluation of PUIs. In doing so, frontline providers will have valuable frameworks of support for identify, isolate, inform approaches^7-10^ in evaluating patients amid rapidly emerging infectious disease outbreaks.

## Data Availability

The data underlying this article cannot be shared publicly due to the privacy of individuals that participated in the study.

## ACKNOWLEDGEMENTS

The authors would like to thank Jasmine B. Ha, MS, Mass General Brigham Digital Health, for project management support of the MGB MPX Digital Health response.

## Accessing .MONKEYPOX

The MPX CDSS tool is available to facilities within the Mass General Brigham (MGB) system for the initial determination of whether a patient meets Person Under Investigation (PUI) criteria for monkeypox (MPX). The tool may be accessed as shown in the following series of figures, which are derived from the chart of a test patient.

### Establishing and detailing PUI criteria

The tool may be accessed by opening any note in Epic and typing “.monkeypox”, as shown in **Supplement, Figure 1**. This will populate the note with the tool, which will guide the user through the relevant questions.

The MPX CDSS tool is designed to proceed through different levels of detail according to the individual patient. The PUI evaluation starts with the assessment of whether the patient has had an otherwise unexplained rash in the last 21 days. An answer may be chosen from one of the dropdown options as shown in **Supplement, Figure 2** and clicking on the appropriate response. If the patient does not have a rash, the tool will indicate that the patient does not meet PUI criteria, making monkeypox unlikely, as shown in **Supplement, Figure 3**.

If the patient meets the rash criterion, a new series of options including both current CDC epidemiological criteria and risk factors relating to the classical transmission of monkeypox will populate (**Supplement, Table 1**). Full text, where this is not visible within the box shown in **Supplement, Figure 4**, can be seen by hovering over each option. Text in the version of the tool as of August 1, 2022 is as follows, and can be readily modified at any time as epidemiological trends dictate.

1. Contact with person(s) with a similar appearing rash.
2. Contact with person(s) with suspected or confirmed MPX.
3. Close/intimate in-person contact with individuals in a social network experiencing MPX activity (e.g., men who have sex with men).
4. Travel to a country with confirmed cases of MPX.
5. Travel to a country where MPX is endemic (Cameroon, Central African Republic, Cote d’Ivoire, Democratic Republic of the Congo, Gabon, Liberia, Nigeria, Republic of the Congo, and Sierra Leone).
6. Contact with a dead/live animal/pet that is an African endemic species or used a product derived from such animals (e.g., game meat, creams, lotions, powders, etc.).
7. No epidemiological risk factors.

If the patient has no epidemiological risk factors as in #7 above, the tool will indicate that the patient does not meet PUI criteria, making monkeypox unlikely, as shown in **Supplementary Figure 5**. Text here, however, includes instructions to call a facility-appropriate backup physician to discuss further if concern for monkeypox on the part of the evaluating clinician remains high for any reason. At MGH, this backup is the Biothreats service, which is staffed at all times by a rotating series of infectious diseases physicians tasked with the initial triage of patients who present with concern for high consequence infectious diseases.

If the patient meets PUI clinical criteria and epidemiological criteria, additional questions will populate according to the epidemiological risk factor chosen. Text used in the current version of the tool is as follows, where *** prevents a note from being signed without the question being answered or the *** otherwise being manually removed:

1. Contact with person(s) with a similar appearing rash.
  a. What was the nature of the contact? ***
  b. When did the contact occur? ***
  c. Where did the contact occur? ***
  d. Did the contact have a rash or other symptoms during the period of contact? Yes. {MPX Symptoms:58856} {MPX Sx Onset Date:58857}
    i. Answering yes to the question of rash or other symptoms during the period of contact will prompt additional boxes specifying the symptoms and date of onset as shown in **Supplement, Figure 6**.
2. Contact with person(s) with suspected or confirmed MPX.
  a. How was the patient notified their contact has suspected or confirmed MPX? ***
  b. What was the nature of the contact? ***
  c. When did the contact occur? ***
  d. Where did the contact occur? ***
  e. Did the contact have a rash or other symptoms during the period of contact? Yes. {MPX Symptoms:58856} {MPX Sx Onset Date:58857}
    i. Answering yes to the question of rash or other symptoms during the period of contact will prompt additional boxes specifying the symptoms and date of onset as in **Supplement, Figure 6** above.
3. Close/intimate in-person contact with individuals in a social network experiencing MPX activity (e.g., men who have sex with men).
  a. Additional questions populating from an affirmative answer to this question are as in #1 above.
4. Travel to a country with confirmed cases of MPX.
  a. To which countries did the patient travel? ***
  b. From what date to what date was the patient in which countries? ***
  c. What was the patient doing in each of their travel destinations? ***
5. Travel to a country where MPX is endemic (Cameroon, Central African Republic, Cote d’Ivoire, Democratic Republic of the Congo, Gabon, Liberia, Nigeria, Republic of the Congo, and Sierra Leone).
  a. Additional questions populating from an affirmative answer to this question are as in #4 above.
6. Contact with a dead/live animal/pet that is an African endemic species or used a product derived from such animals (e.g., game meat, creams, lotions, powders, etc.).
  a. What was the nature of the animal contact or contact with products? ***
7. No epidemiological risk factors.

### Immediate next steps

After detailing the specific questions related to a patient’s epidemiological risk, the clinician will be presented with text specifying immediate next steps, including isolation guidelines appropriate to the clinician’s location. An example is shown in **Supplement, Figure 7**.

### Obtain additional information

After reviewing immediate next steps, the clinician will be presented with a series of additional questions detailing the patient’s clinical history, specifically the timing, duration, and location of the patient’s rash. Additional boxes will prompt the identification of any other symptoms present and when these started. An example is shown in **Supplement, Figure 8**.

After answering clinical questions as above, the tool will prompt the clinician to answer a series of questions relating to the safe disposition of the patient, which are to aid in decision making on the part of the physician approving testing – at MGH, the Biothreats service – on whether the patient can safely isolate at home. These are shown in **Supplement, Figure 9**. Questions on whether the patient lives alone or has pets will prompt additional questions as shown in **Supplement, Figures 10** and **11**.

### Obtain photos

Text will prompt the clinician evaluating the patient (in person or virtually) to upload photos of the rash in question to the chart. This may assist in assessment of the risk that the patient has monkeypox as well as decisions on whether to pursue therapy with tecovirimat under an investigational protocol. Specific language under this section may be seen in **Supplement, Figures 10** and **11**.

## Limitations

Although the use of the CDS tool enables the efficient triage of many patients who may meet PUI criteria, clinical judgment is still required for the appropriate management of individual patients. In general, the CDSS is written to be broad – more sensitive than specific – in the hope that this will maintain a low threshold for the identification of patients who may have monkeypox. Further review may then be obtained from a backup physician who can decide what, if any, additional evaluation needs to be done. At MGH, this backup is provided by a dedicated Biothreats service, as discussed above, though infectious disease specialists may serve in a similar capacity at many institutions without the benefit of a separate service.

Conversely, we have found that there are instances in which the evaluating clinician retains a high suspicion for monkeypox despite the patient not meeting PUI criteria. In these instances, the tool directs the clinician to discuss further with a backup physician as above, who is empowered to override the tool should it be deemed clinically appropriate.

## Summary

We have described here the implementation of a CDSS that enables frontline clinicians to efficiently determine whether a given patient meets PUI criteria for monkeypox. When this is the case, the tool elicits further information that facilitates additional decisions in consultation with a backup physician service on questions of testing, isolation, and potentially treatment. Importantly, the MPX CDSS Tool is simple to adapt to changing conditions in an outbreak, as has already been done several times in the first 6 weeks of implementation. Thus, care is streamlined by allowing a small subset of physicians to stay fully abreast of an outbreak and, through the centralized maintenance CDSS, promote best practices consistent with the most recent data available.

## Further Information

For further information on the MPX CDSS Tool and potential for adapting similar strategies at different institutions, please contact David M. Rubins, MD

**Supplement Table 1.** CDC Person Under Investigation Criteria for Monkeypox, adapted for Mass General Brigham system, 2022.

|  |  |
| --- | --- |
| **Epidemiological Criteria (one of the following) within 21 days of symptom onset** | 1. Reports having contact with a person or people with a similar appearing rash or who received a diagnosis of confirmed or probable monkeypox **or**<br>2. Had close or intimate in-person contact with individuals in a social network experiencing monkeypox activity, this includes men who have sex with men (MSM) who meet partners through an online website, digital application (“app”), or social event (e.g., a bar or party)<br>3. Traveled outside the US to a country with confirmed cases of monkeypox or where MPXV is endemic (Cameroon, Central African Republic, Cote d’Ivoire, Democratic Republic of the Congo, Gabon, Liberia, Nigeria, Republic of the Congo, and Sierra Leone), **or**<br>4. Had contact with a dead or live wild animal or exotic pet that is an African endemic species or used a product derived from such animals (e.g., game meat, creams, lotions, powders, etc.) |
| **Clinical Criteria** | A new and otherwise unexplained skin rash (exhibiting macular, papular, vesicular, or pustular lesions; generalized or localized; discrete or confluent). The CDC provides images that can assist with clinical recognition here.<br>*Note that patients may present with a flu-like illness (e.g., fever, chills, malaise, headache, muscle aches) prodrome and new lymphadenopathy (periauricular, axillary, cervical, or inguinal). Illness could be clinically confused with a sexually transmitted infection like syphilis or herpes, or with varicella zoster virus.* |

Sources: CDC Monkeypox, 2022 and MDPH Monkeypox, 2022.

**Supplementary Figure 1:**
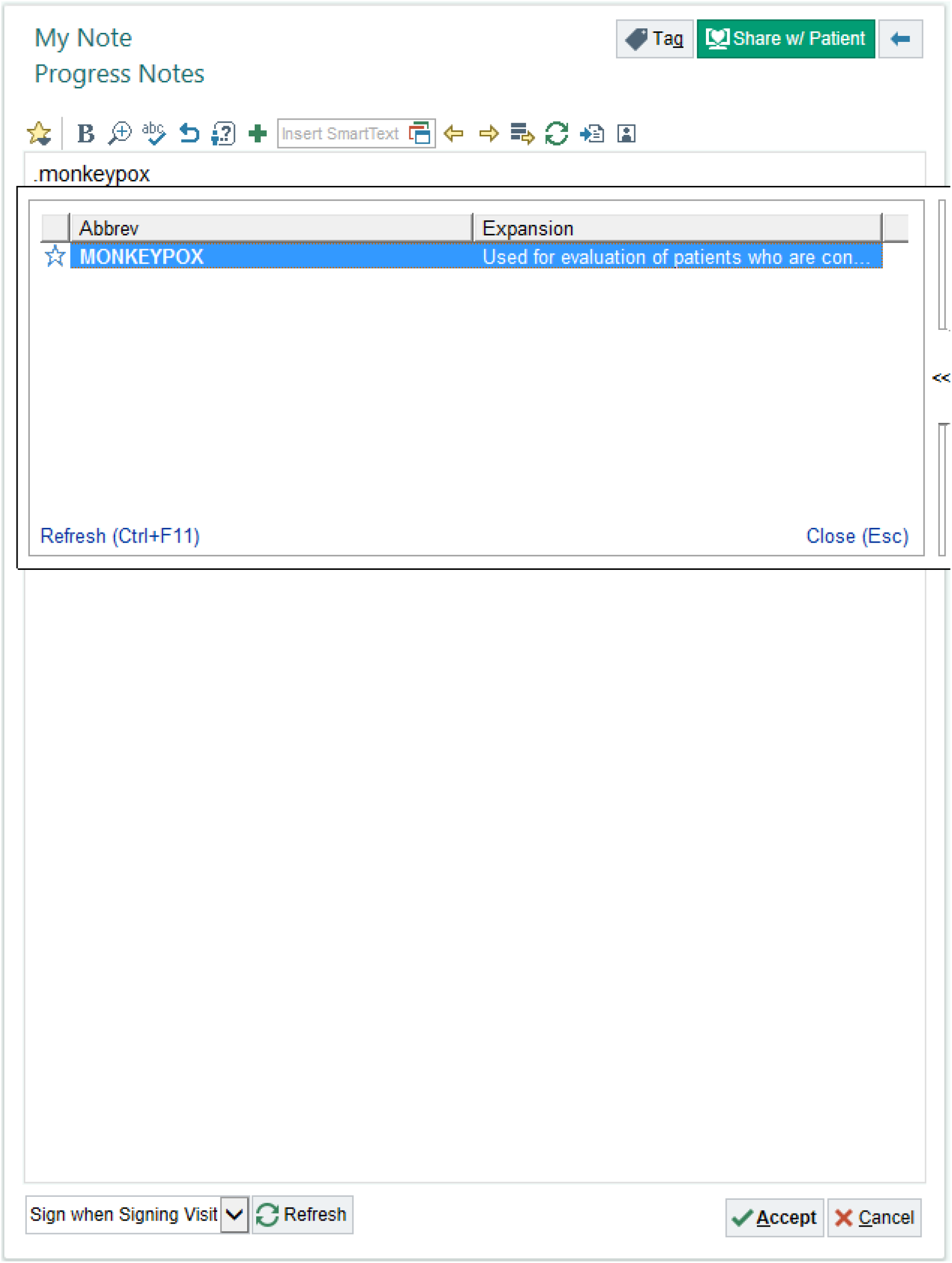
Initial appearance of the .monkeypox SmartLink.

**Supplementary Figure 2:**
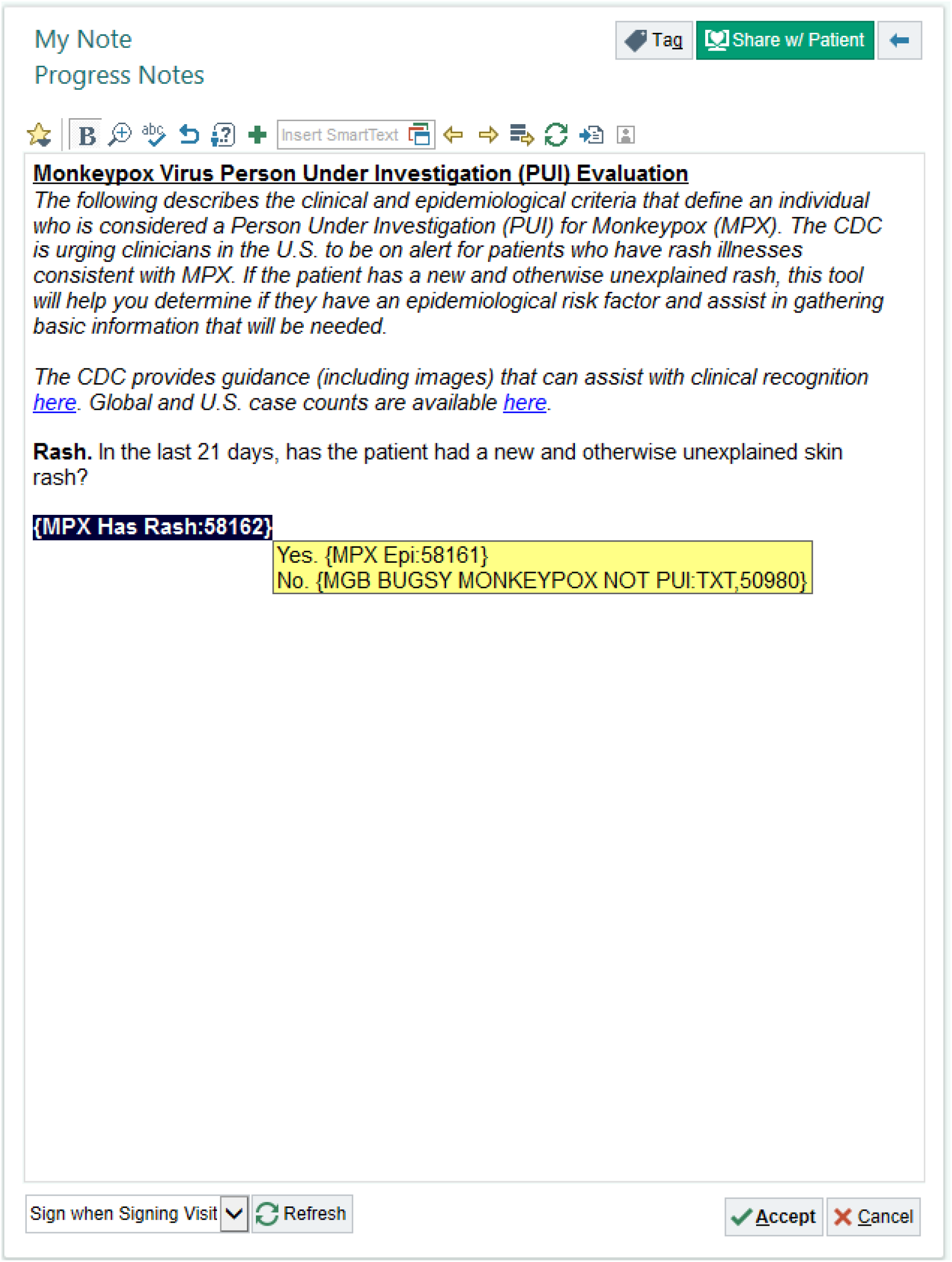
Rash evaluation in the SmartLink.

**Supplementary Figure 3:**
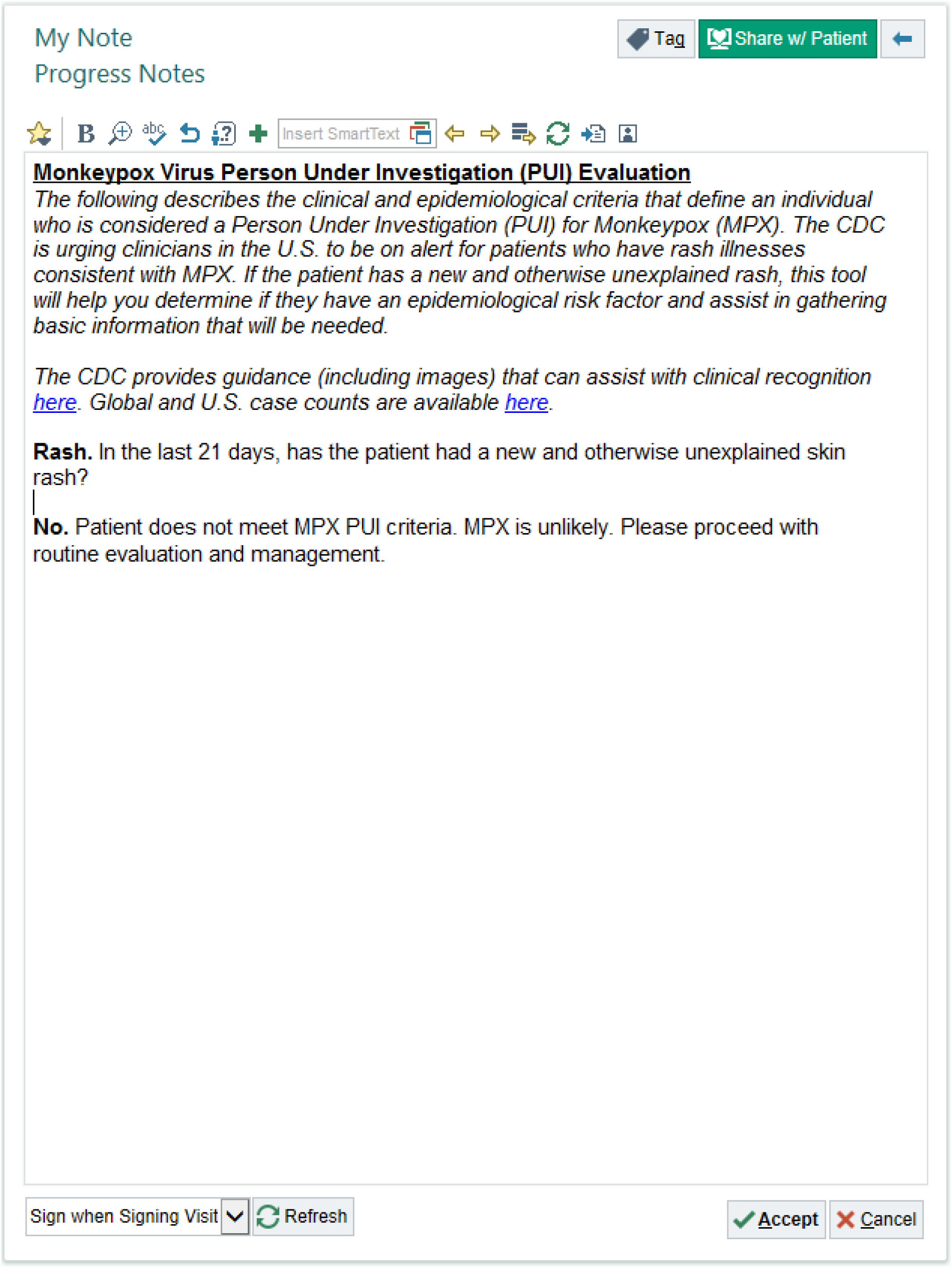
Text generated in the event that patient fails to meet PUI clinical criteria.

**Supplementary Figure 4:**
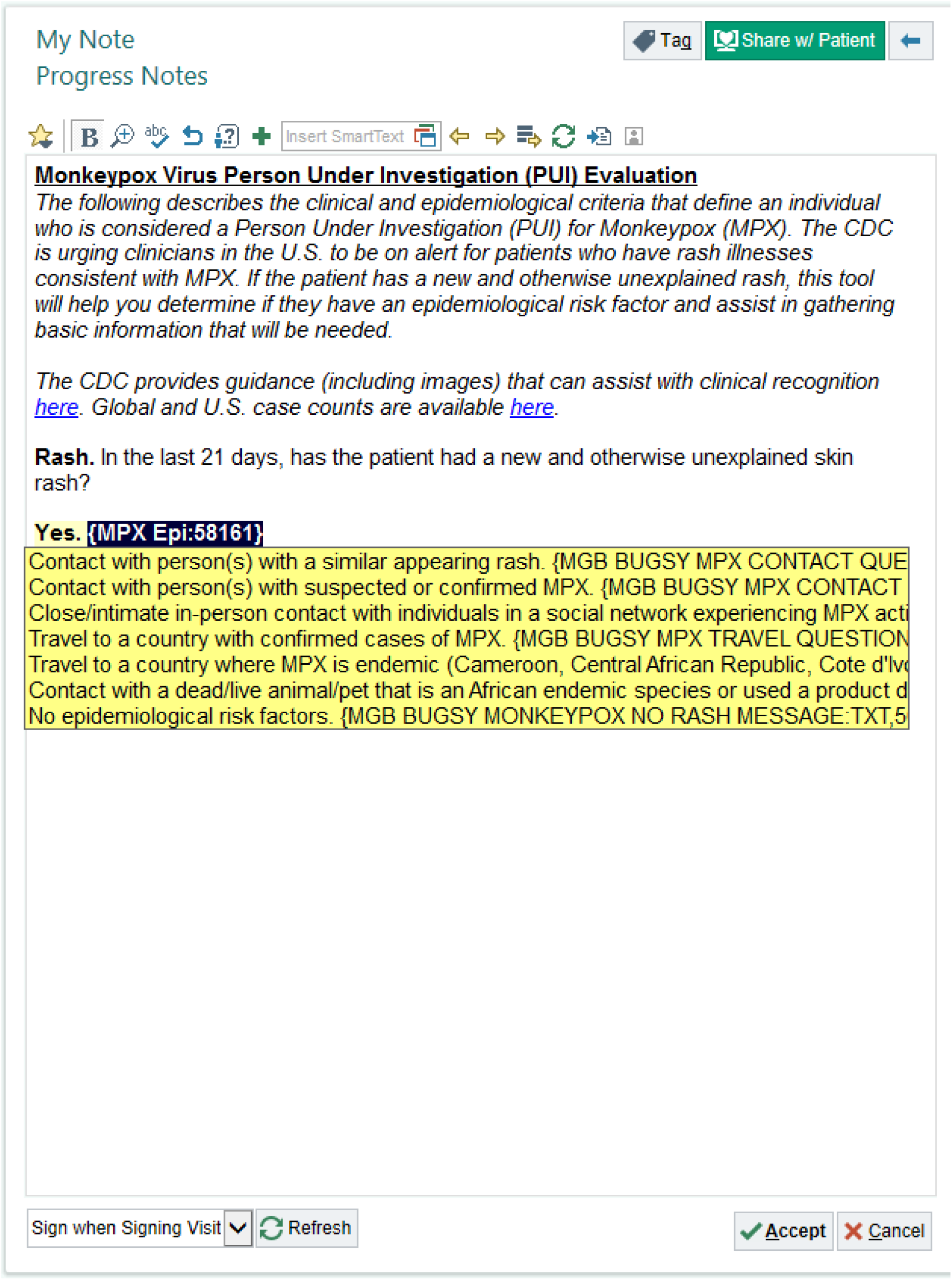
Evaluation of epidemiological risk factors.

**Supplementary Figure 5:**
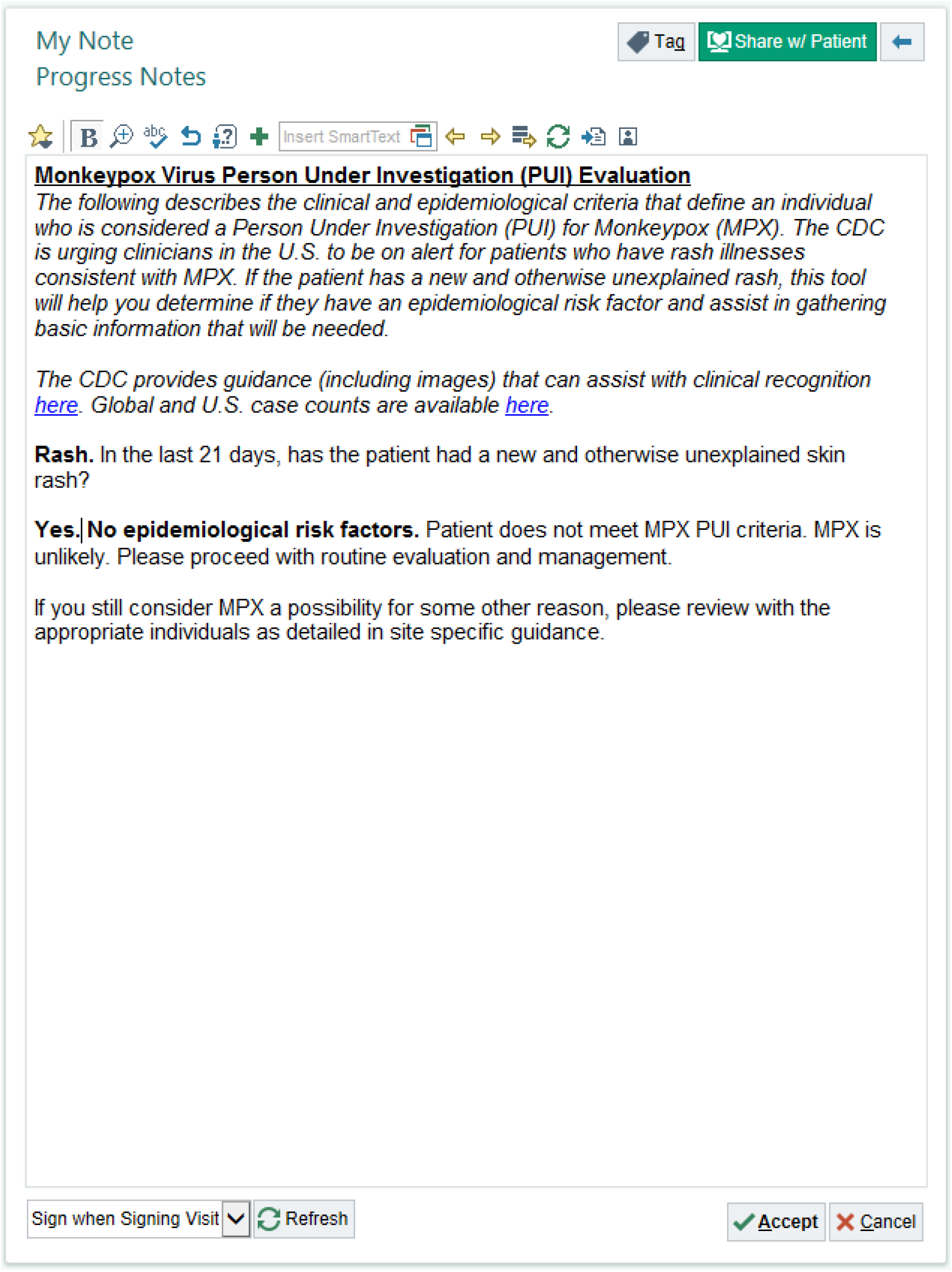
Text generated in the event that patient fails to meet PUI epidemiological criteria.

**Supplementary Figure 6:**
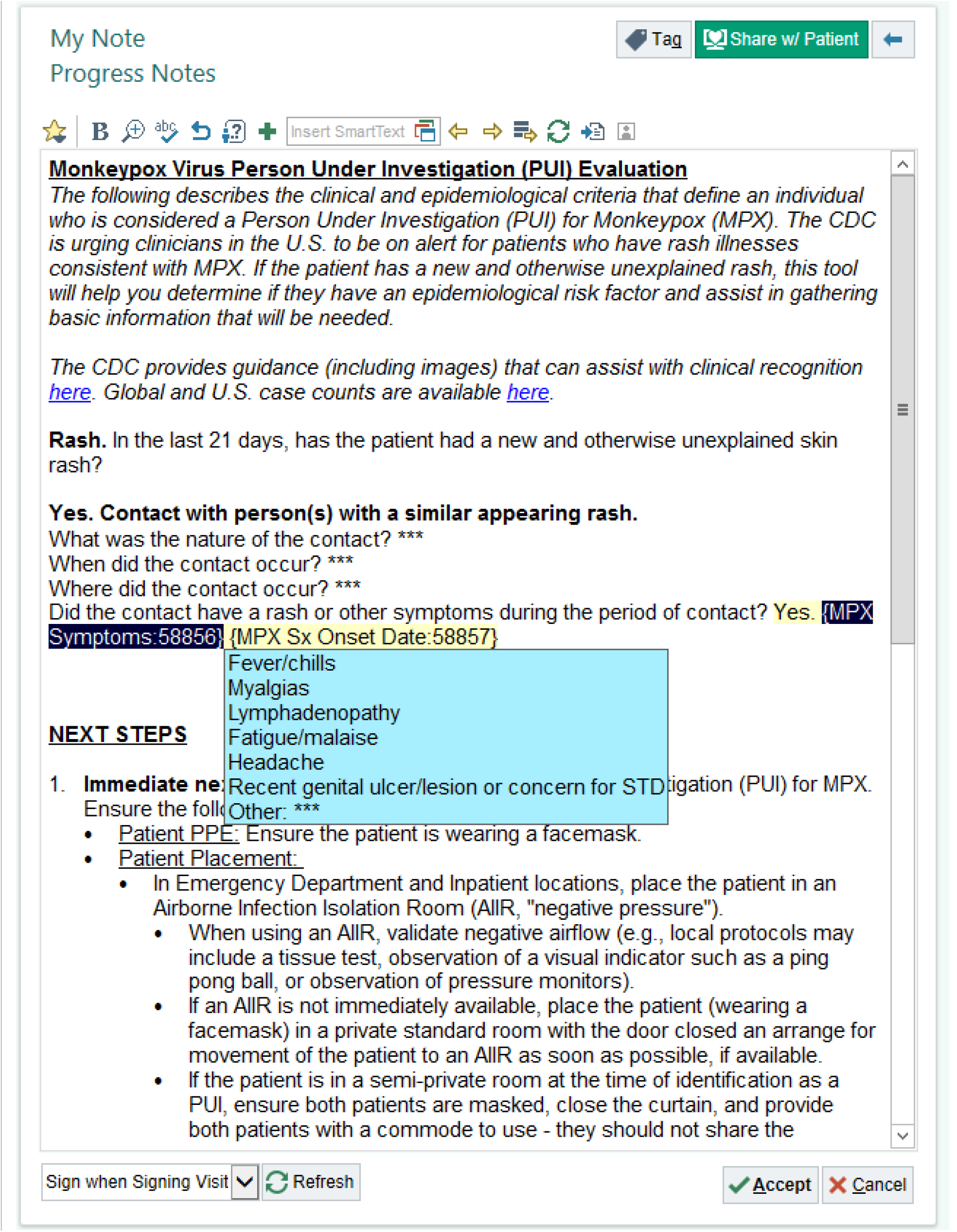
Detailed questions about symptoms in a patient’s contact at the time of contact.

**Supplementary Figure 7:**
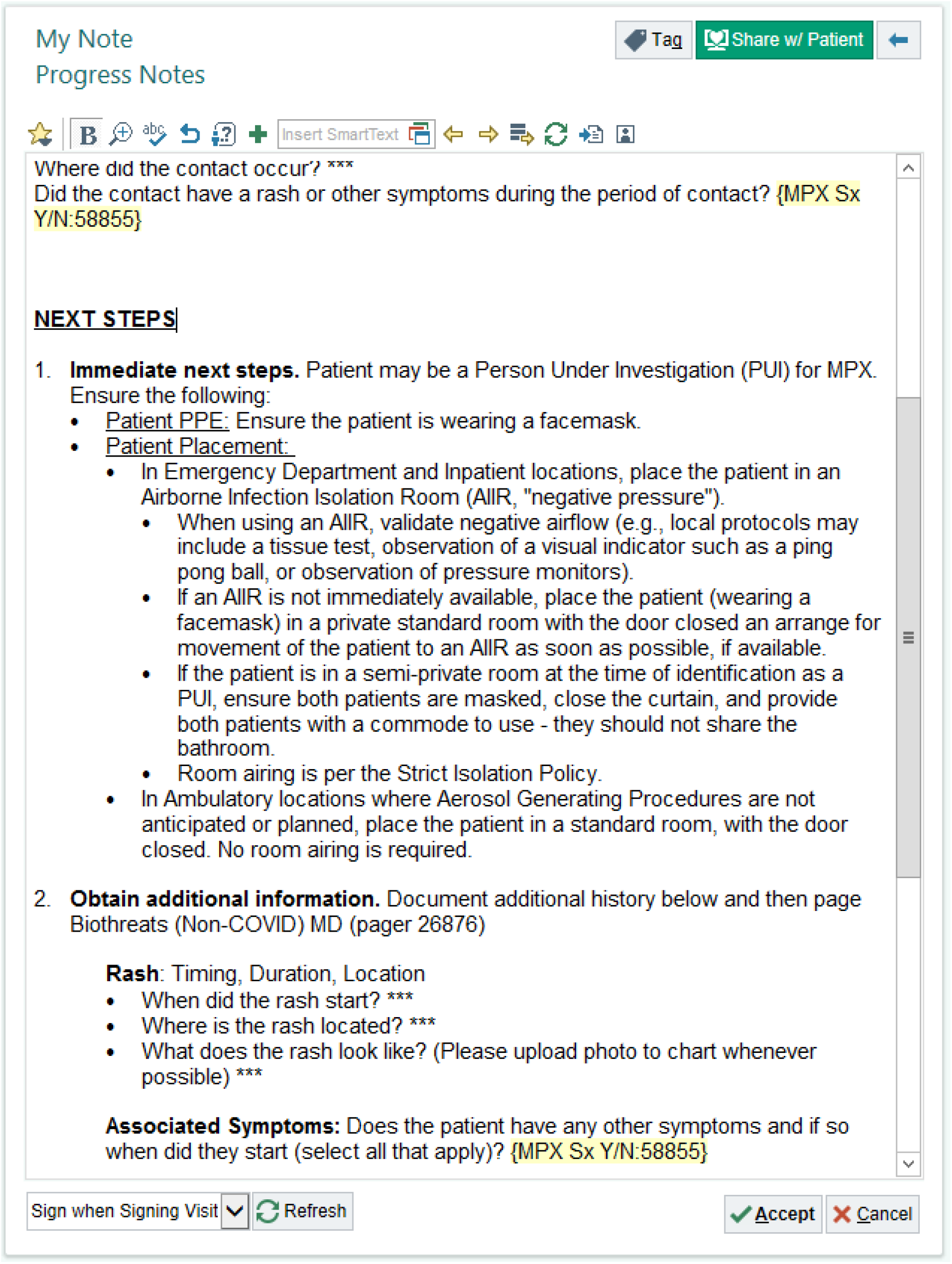
Guidance on immediate next steps in the management of patients who meet PUI criteria.

**Supplementary Figure 8:**
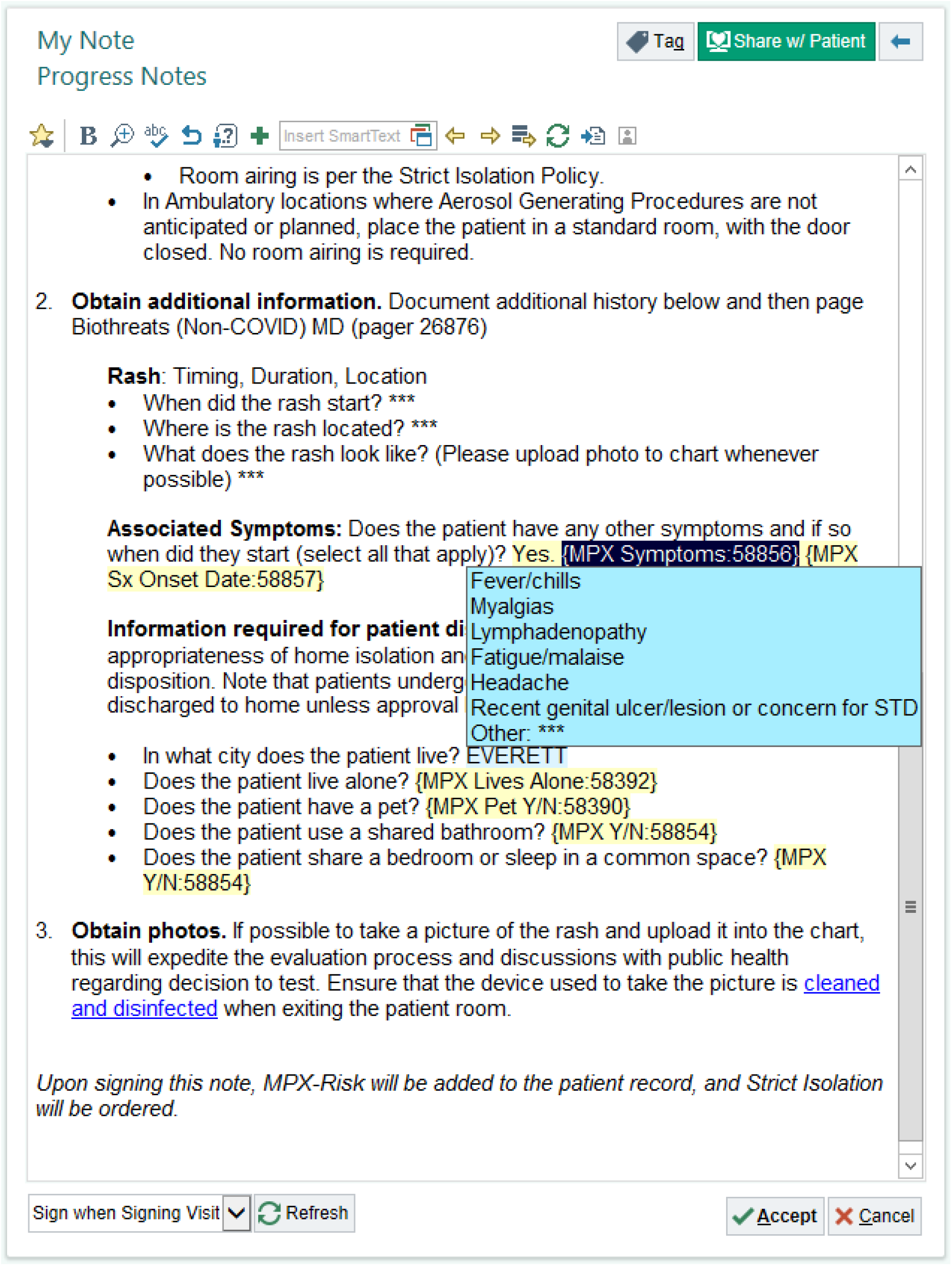
Additional clinical questions once a patient has been identified as a potential PUI.

**Supplementary Figure 9:**
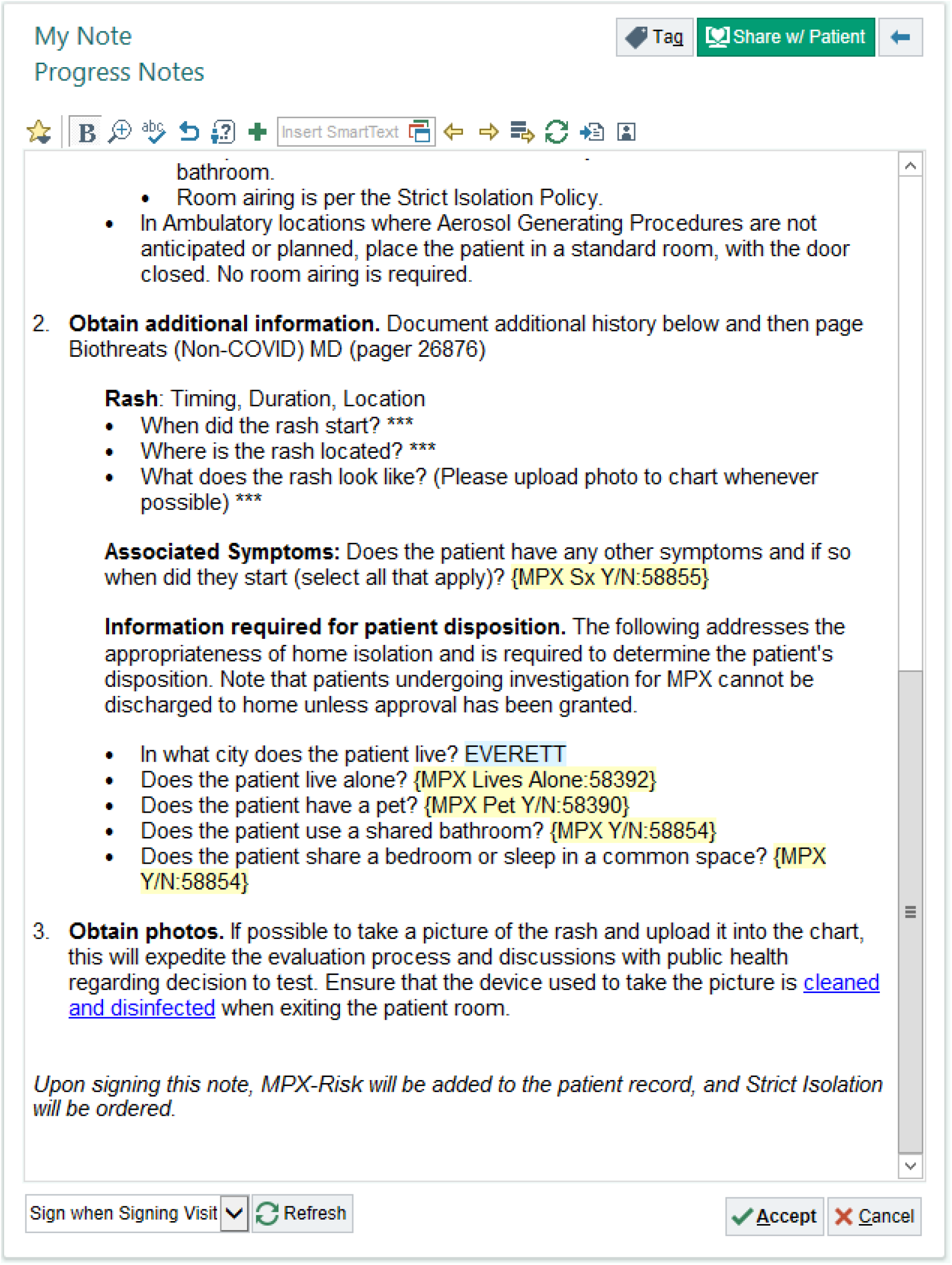
Questions pertinent to the safe disposition of patients who will isolate at home pending monkeypox test results.

**Supplementary Figure 10:**
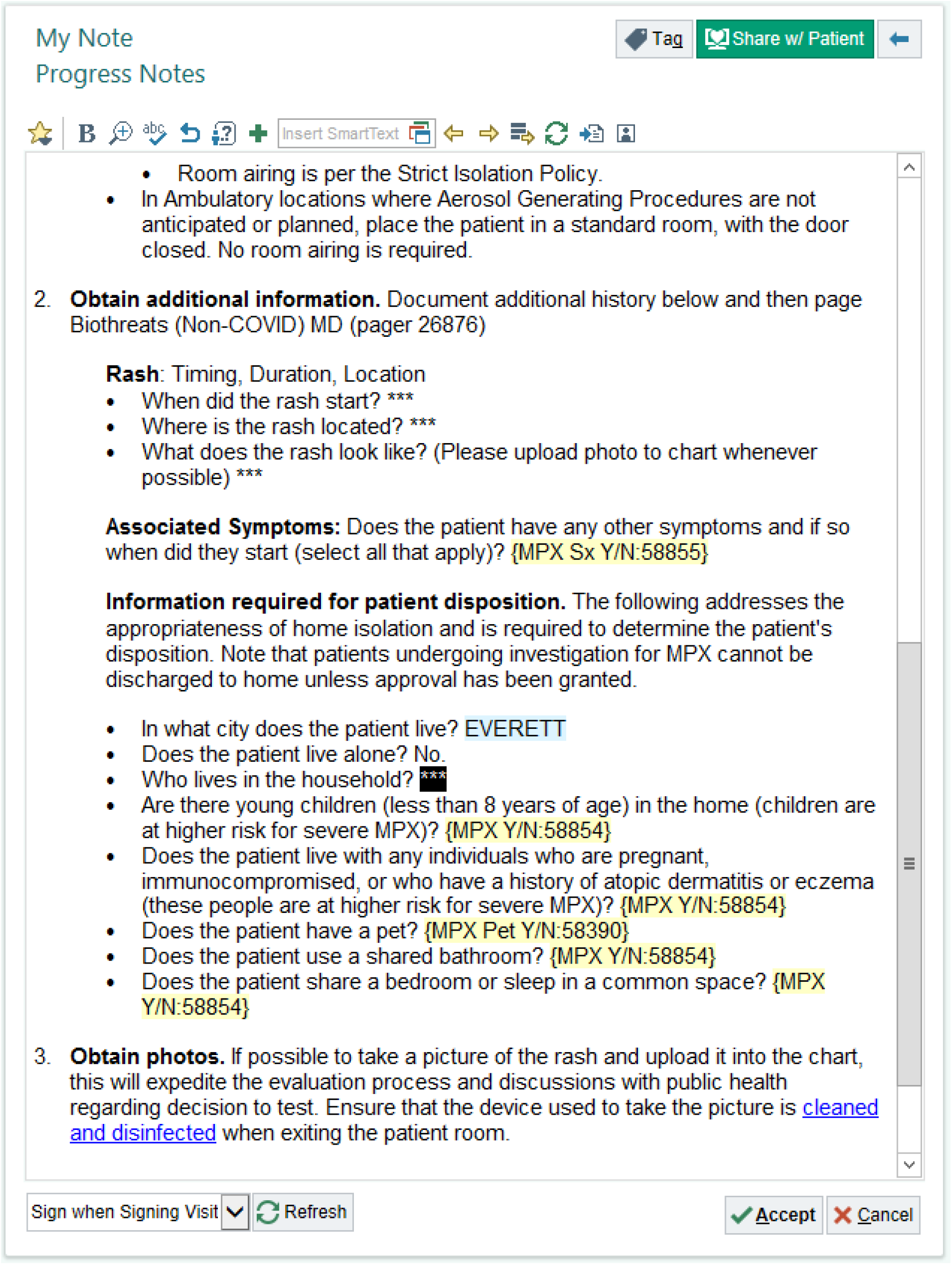
Questions about individuals with whom the patient lives in the event that the patient does not live alone.

**Supplementary Figure 11:**
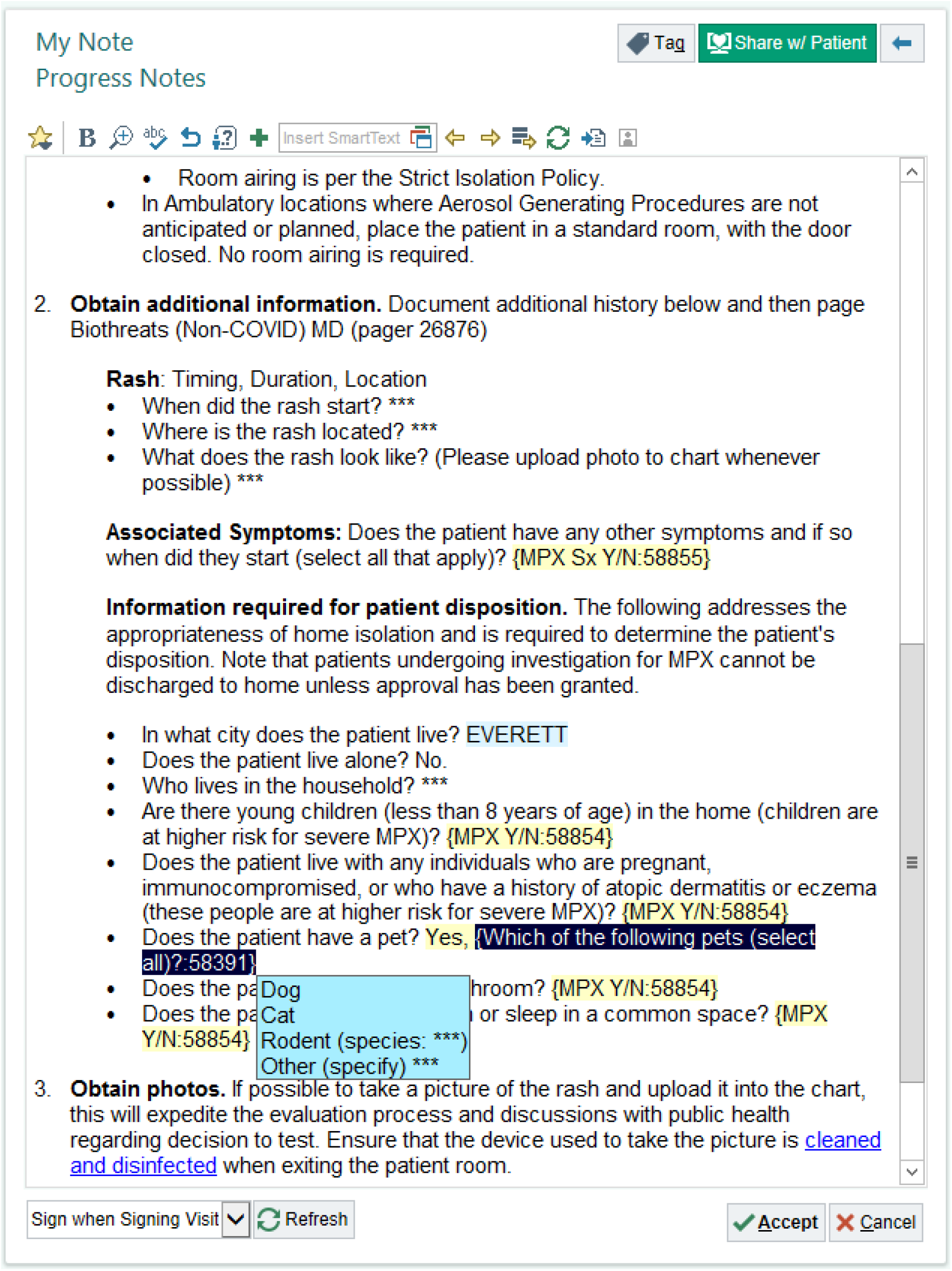
Questions about any pets that may be in the patient’s living environment.

