## Supplementary figures and images for "Development and implementation of a clinical decision support system tool for the evaluation of suspected monkeypox infection"

### Figure 1

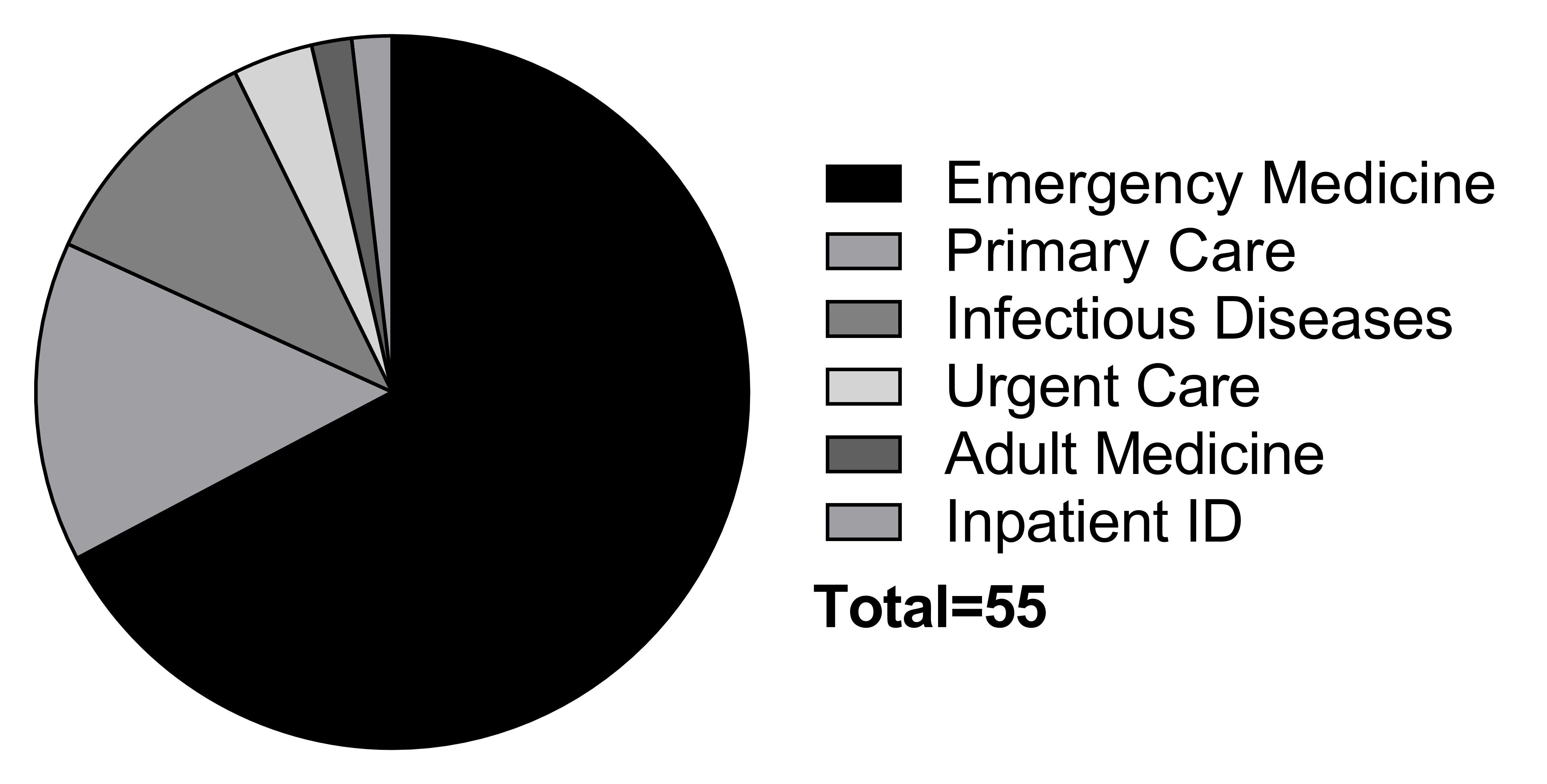

### Figure 2

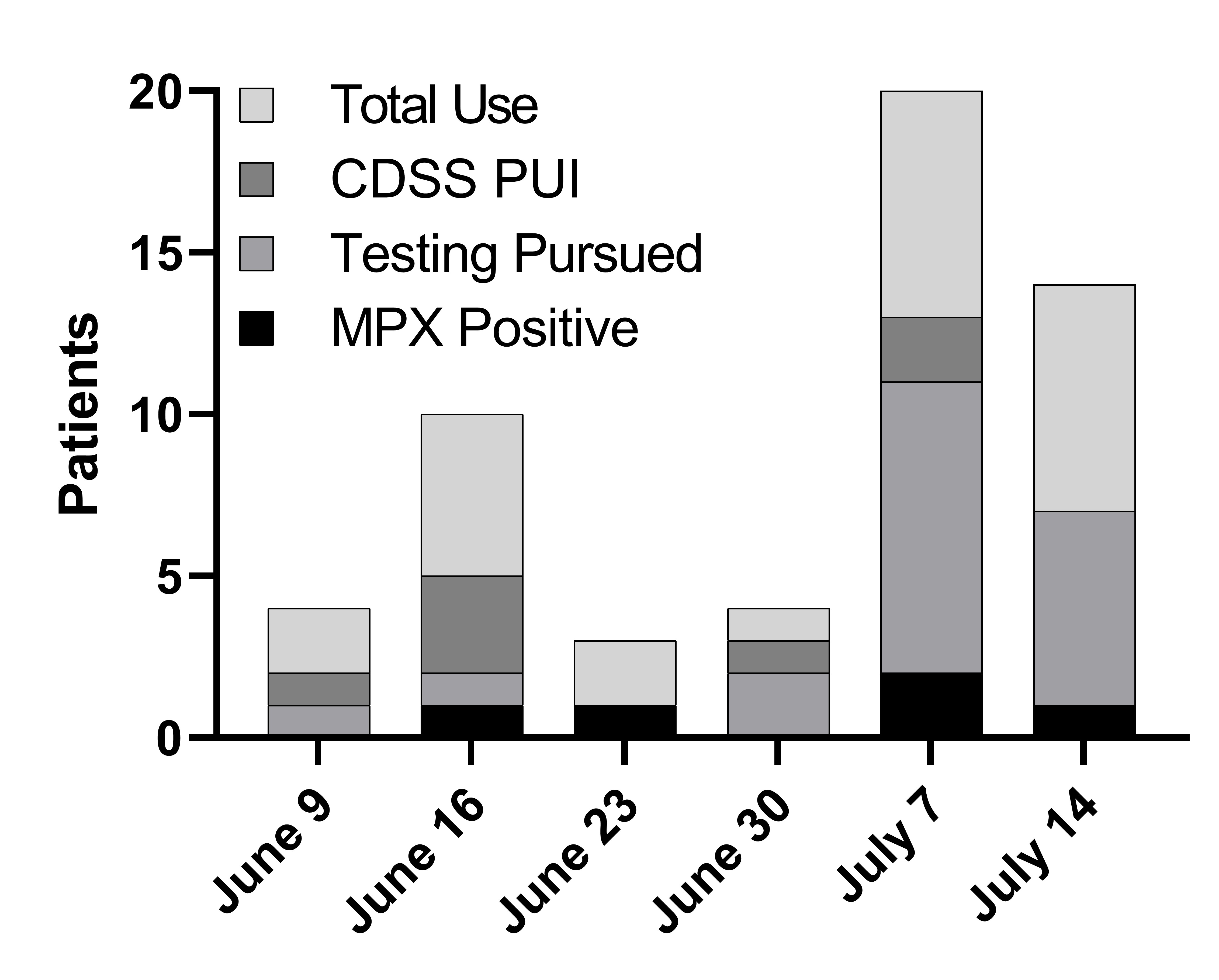

### Figure 3

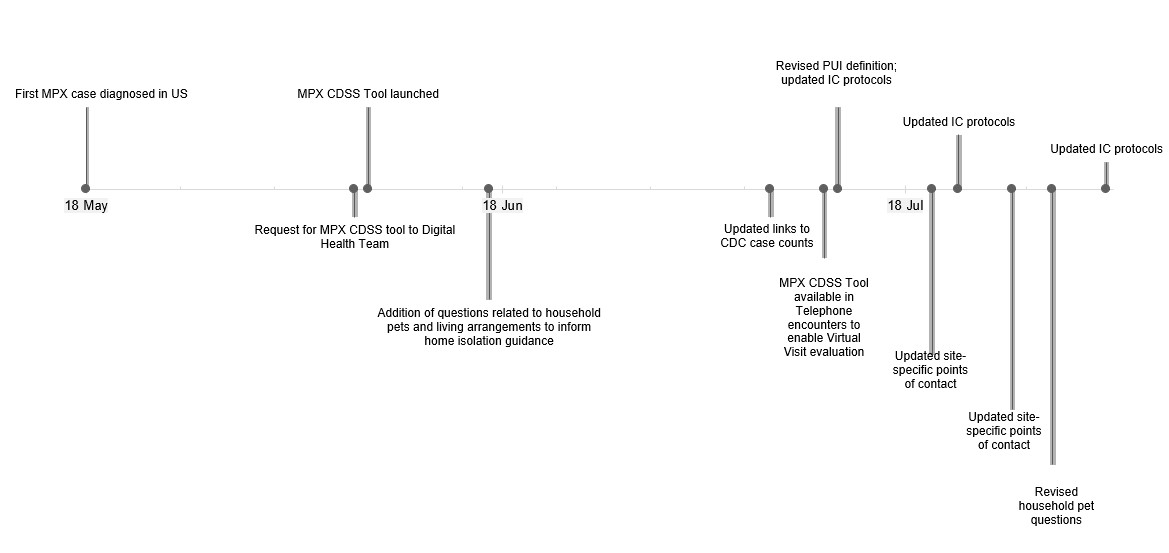
